# The Influence of Time-Limited Immunity on a COVID-19 Epidemic: A Simulation Study

**DOI:** 10.1101/2020.06.28.20142141

**Authors:** Robert J. Kosinski

## Abstract

A series of spreadsheet simulations using SEIS, SEIR, and SEIRS models showed that different durations of effective immunity could have important consequences for the prevalence of an epidemic disease with COVID-19 characteristics. Immunity that lasted four weeks, twelve weeks, six months, one year, and two years was tested with pathogen R_0_ values of 1.5, 2.3, and 3.0. Shorter durations of immunity resulted in oscillations in disease prevalence. Immunity that lasted from three months to two years produced recurrent disease outbreaks triggered by the expiration of immunity. If immunity “faded out” gradually instead of persisting at full effectiveness to the end of the immune period, the recurrent outbreaks became more frequent. The duration of effective immunity is an important consideration in the epidemiology of a disease like COVID-19.

## Introduction

The ability of the immune system to protect a person recovering from COVID-19 from further infection is an important question in management of the COVID-19 pandemic. The majority of infected individuals do clear the SARS-CoV-2 virus from their bodies, and do create antibodies and T-cells against the virus (reviewed in Prompetchara *et al*, 2020), but how long this response remains protective is unclear. The basic question, “If I’ve had it, can I get it again?” still has no definite answer. The duration of immunity to SARS-CoV-2 is also of great importance for the practicality of a vaccine.

Kissler *et al*. (2020) did a simulation study that explored the duration of immunity plus several other variables such as the strength of social distancing, cross-immunity to flu strains, and seasonal changes in the basic reproduction number (R_0_) to simulate the “postpandemic period” for COVID-19. They found that shorter duration of immunity (about 40 weeks) tended to trigger annual outbreaks, but longer duration (up to two years) produced biennial outbreaks. They found that if immunity was permanent, the disease could disappear after its initial outbreak. These findings were influenced by the fact that Kissler *et al*. increased R_0_ in the fall and decreased it in the spring of each simulated year to model seasonal effects common in flu epidemics. Britton *et al*. (2020) did extensive simulations of a COVID-19-like disease in order to determine the effect of population heterogeneity on the development of herd immunity, but in their models, immunity was assumed to last “for an extended period of time,” longer than the events they simulated.

This report explores the effects of the duration of immunity in a constant, non-seasonal environment. It uses simulation to determine the effect of immune responses that are protective for time periods ranging from four weeks to two years after the conclusion of the infective period.

### Methods--The Model

The simulations were standard SEIS, SEIR, and SEIRS models done using difference equations on an Excel spreadsheet. The iteration interval (each line and column on the spreadsheet) was one day. The most complex model (SEIRS) contained four different groups of individuals in a model population of 1000: susceptible, exposed (infected but not yet infective), infective, and immune, with the following durations:

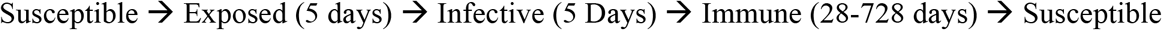

Each exposed, infective, and immune individual moved one day at a time through its group. In the SEIS model, an infective individual cycled back to the susceptible group when its time in the infective group finished; in the SEIR model they permanently entered the immune group when the infection was over; in the SEIRS model, they cycled back to susceptible when their time in the immune group was up.

Although “Exposed” and “Infective” groups were handled separately in the calculations, they were lumped into one group called “Infected” in the graphs that follow.

There was no death or seasonal change in the model. The simulations started when one individual at the very beginning of its five-day infective career entered the population of 999 susceptible individuals.

The number of new infections *per infective* on day t was defined by

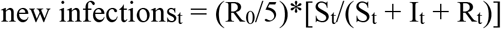

where S_t_, I_t_, and R_t_ are the numbers of susceptible, infected, and immune hosts and the “5” refers to the 5-day length of the infective period.

Because the dynamics varied greatly depending on the transmissibility of the virus, all simulations were done with three values of the basic reproduction number (R_0_): 3.0, 2.3, and 1.5.

One final condition tested was “immune fading,” in which immunity slowly decreases over time rather than lasting undiminished to the end of the immune period. The loss of immunity starts slowly, and then accelerates towards the end of the immune period. This is illustrated in Fig. 1 for a cohort of 1000 newly-immune individuals with a maximum duration of immunity of one year:

**Fig. 1.**
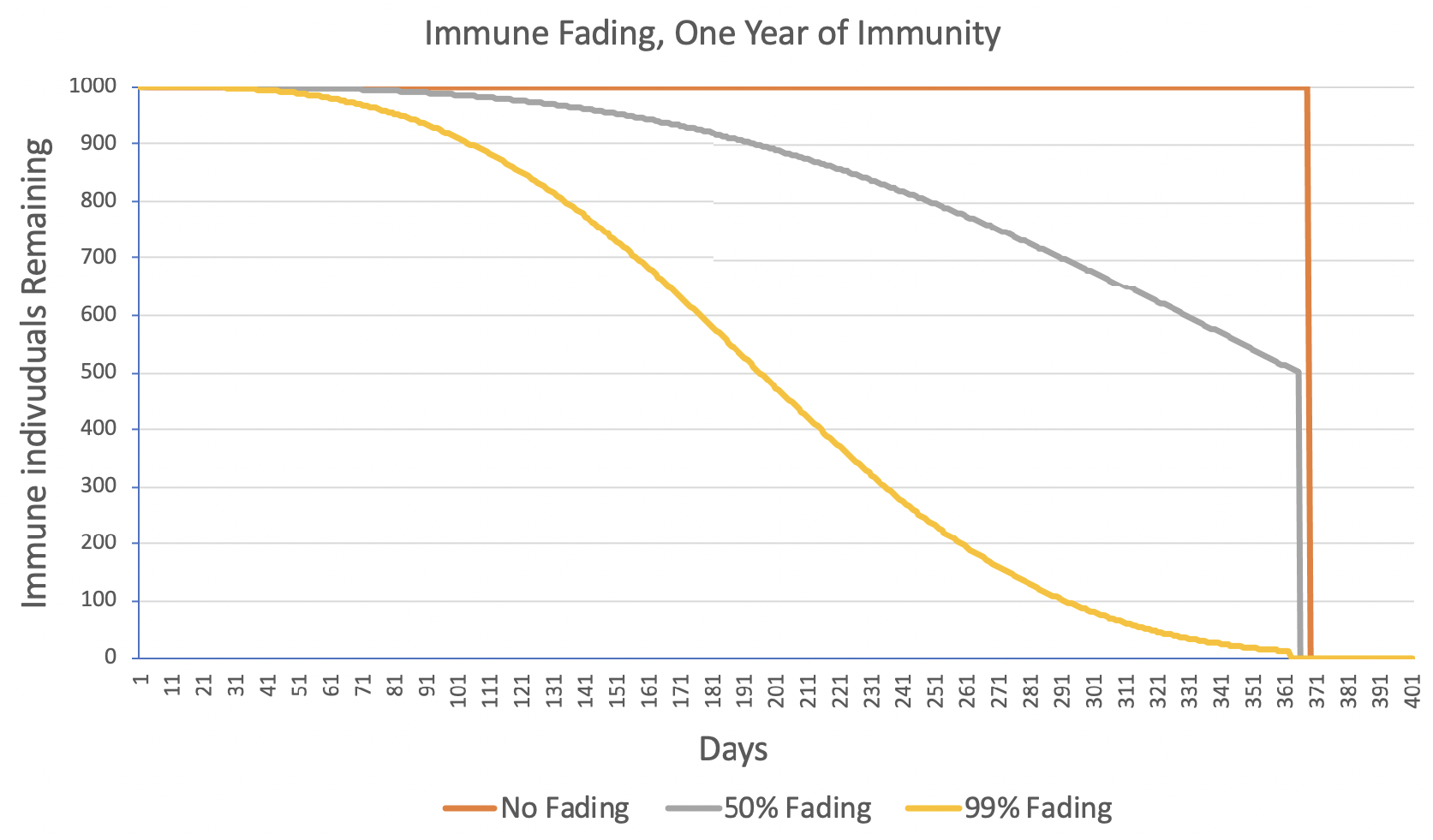
The size of a cohort of 1,000 newly-immune individuals under three conditions of immune “fading.” “50% Fading” means that half the immune individuals lose their immunity before their one year of immunity ended. Maximum duration of immunity was 364 days from the cohort’s origin, after which all the remaining immune individuals become susceptible again.

## Results

The results are shown in Figs. 2-9 below, which extend from no immunity, through immunity of increasing duration, to permanent immunity in Figure 9. Figs. 2-9 do not include any immune fading. Every immune organism experiences its full duration of immunity.

**Fig. 2.**
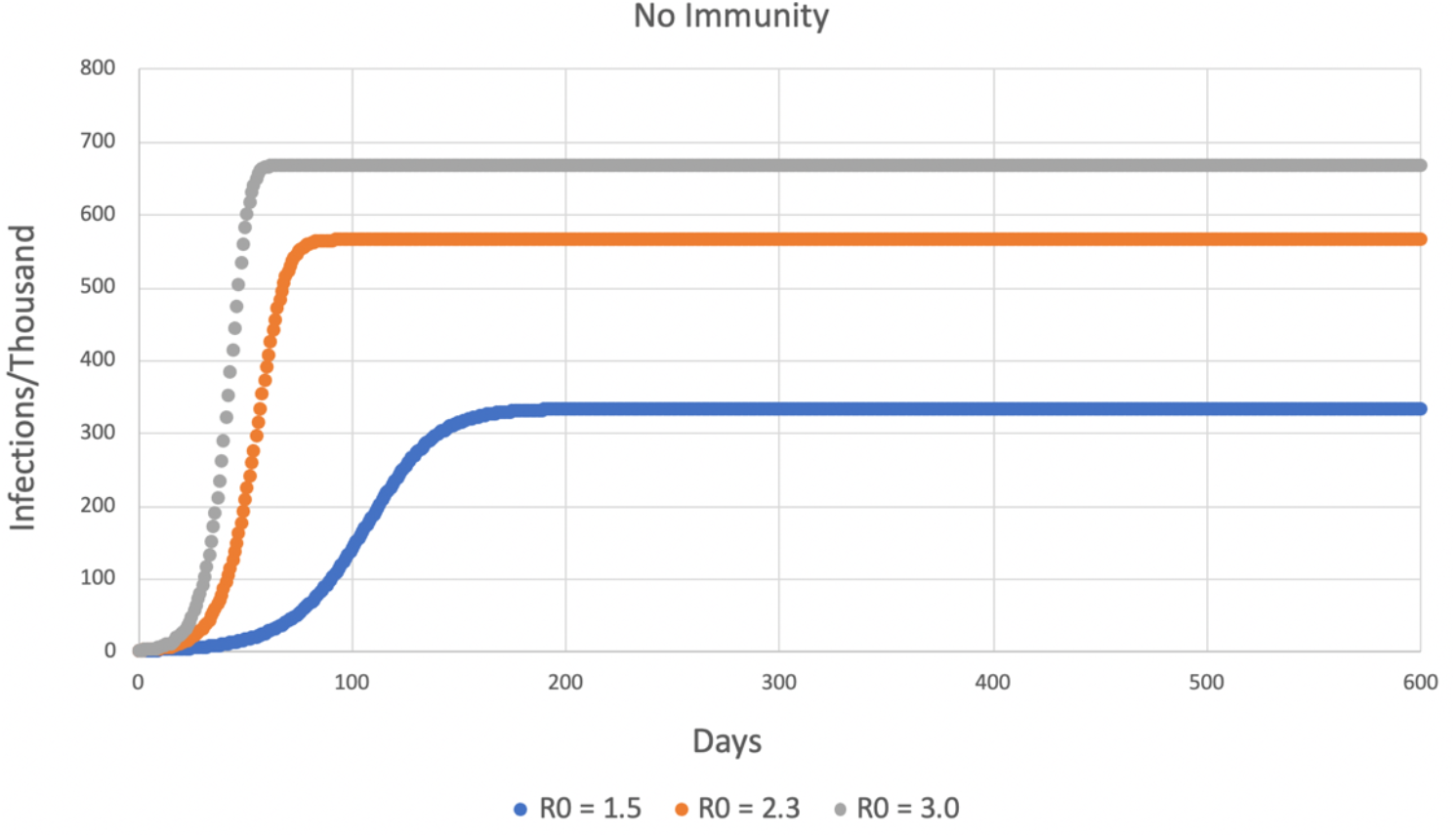
An SEIS model (no immunity). The infections achieve a stable equilibrium where the proportion of infected = 1 - (1/R_0_), as expected. The three curves represent the three tested values of R_0_.

**Fig. 3.**
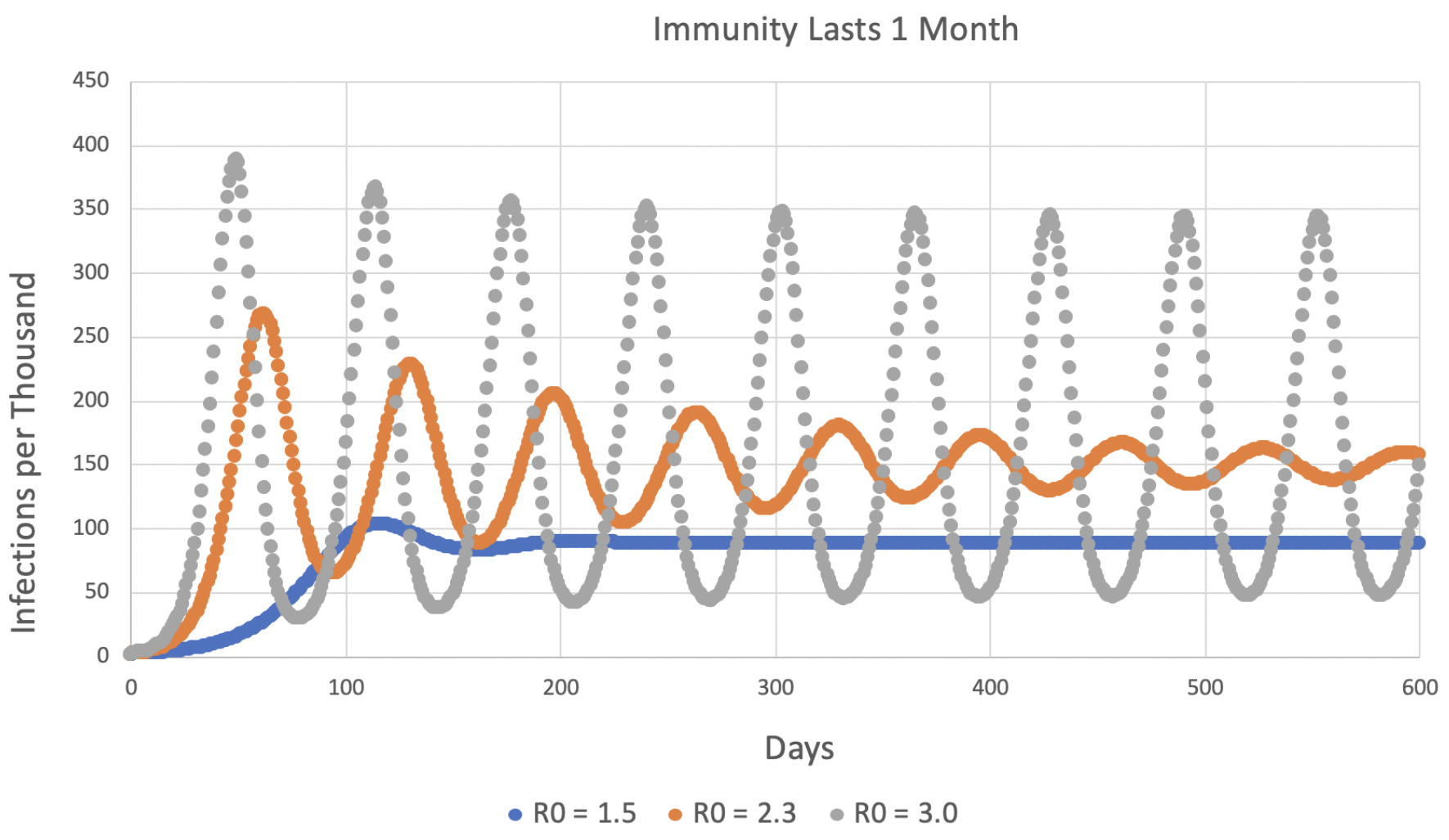
An SEIRS model in which immune individuals return to being susceptible after 28 days of immunity.

**Figure 4.**
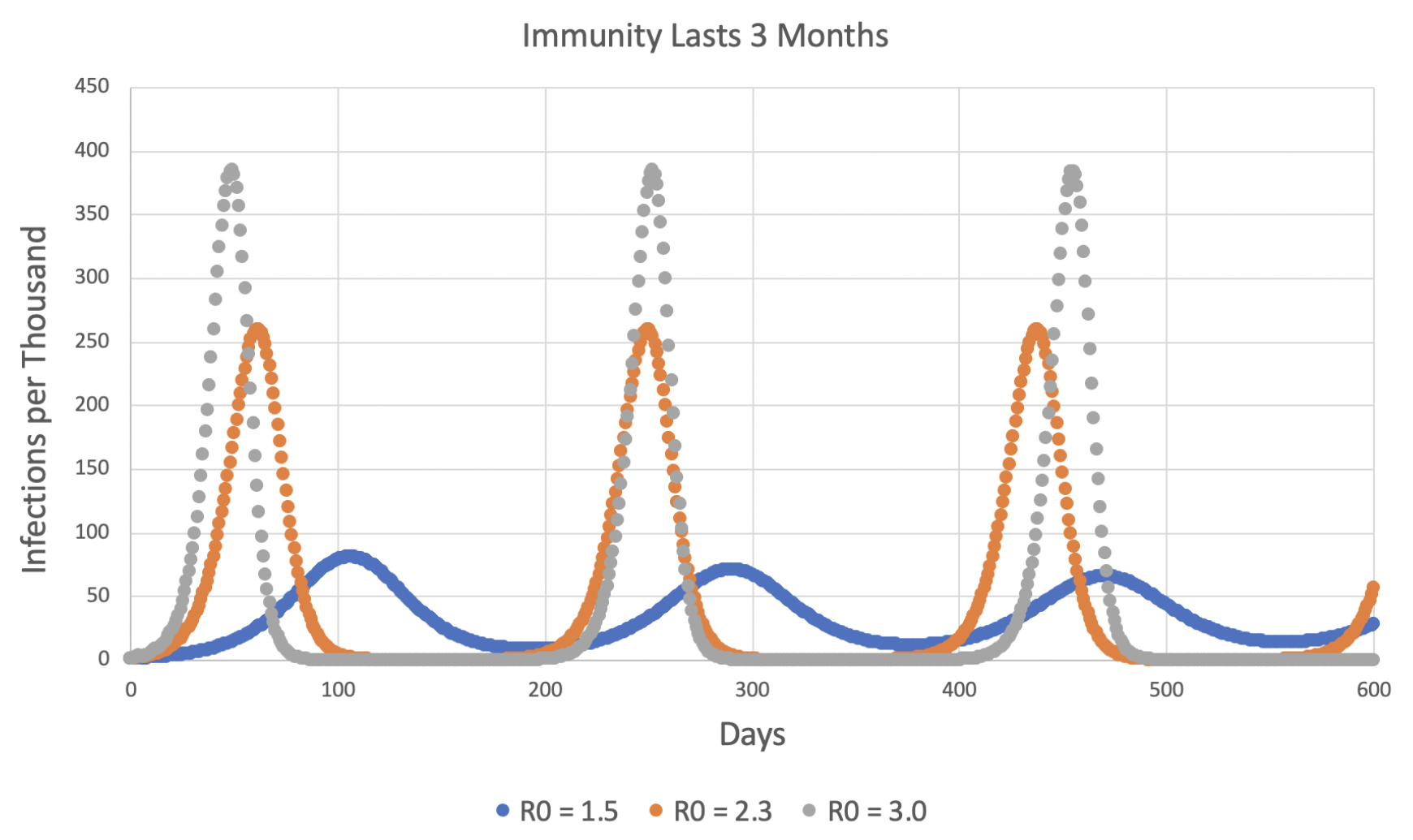
SEIRS model with 12 weeks (3 months) of immunity.

**Figure 5.**
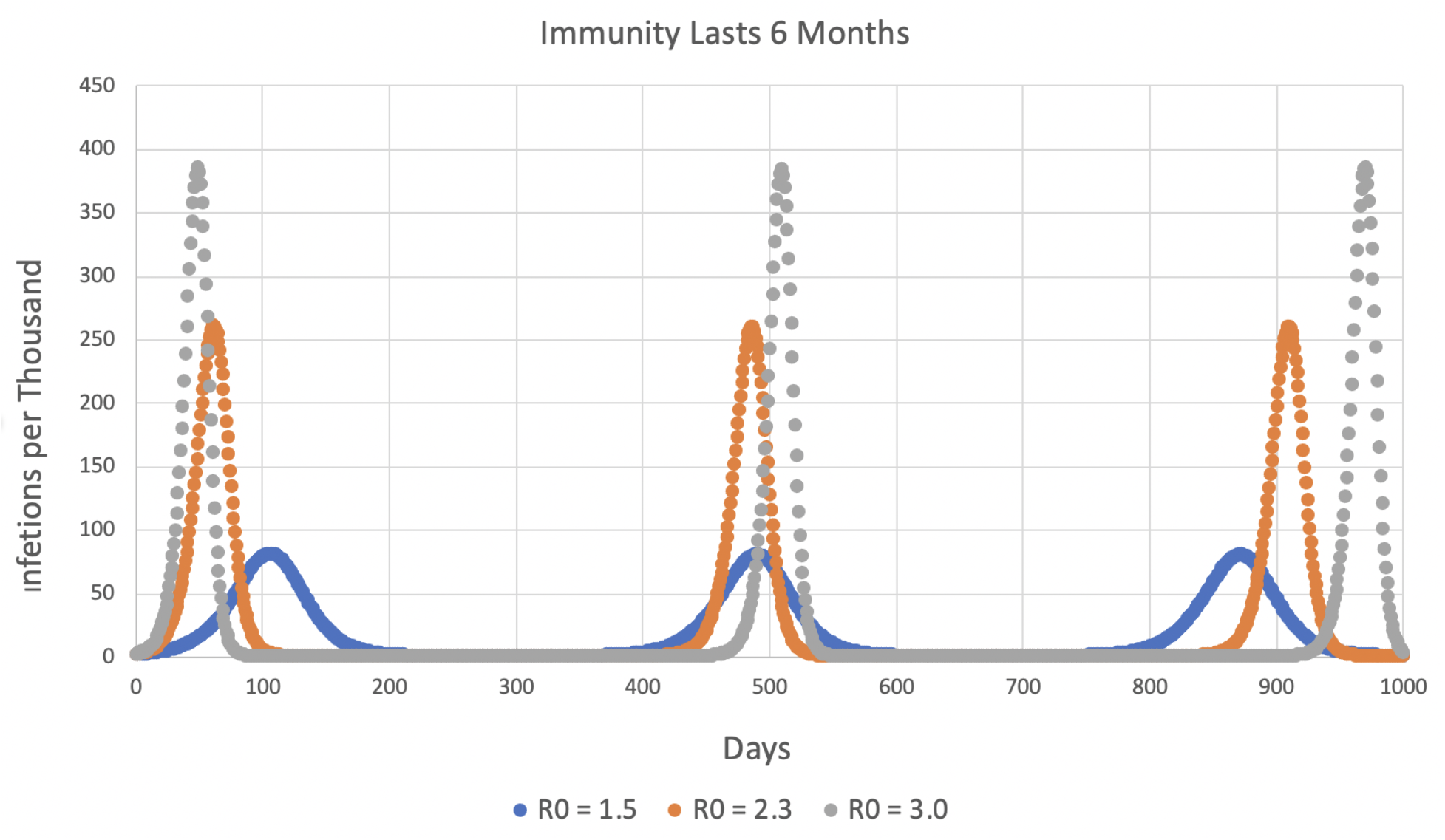
SEIRS model with 26 weeks (6 months) of immunity. X-axis extends to 1000 days.

**Figure 6.**
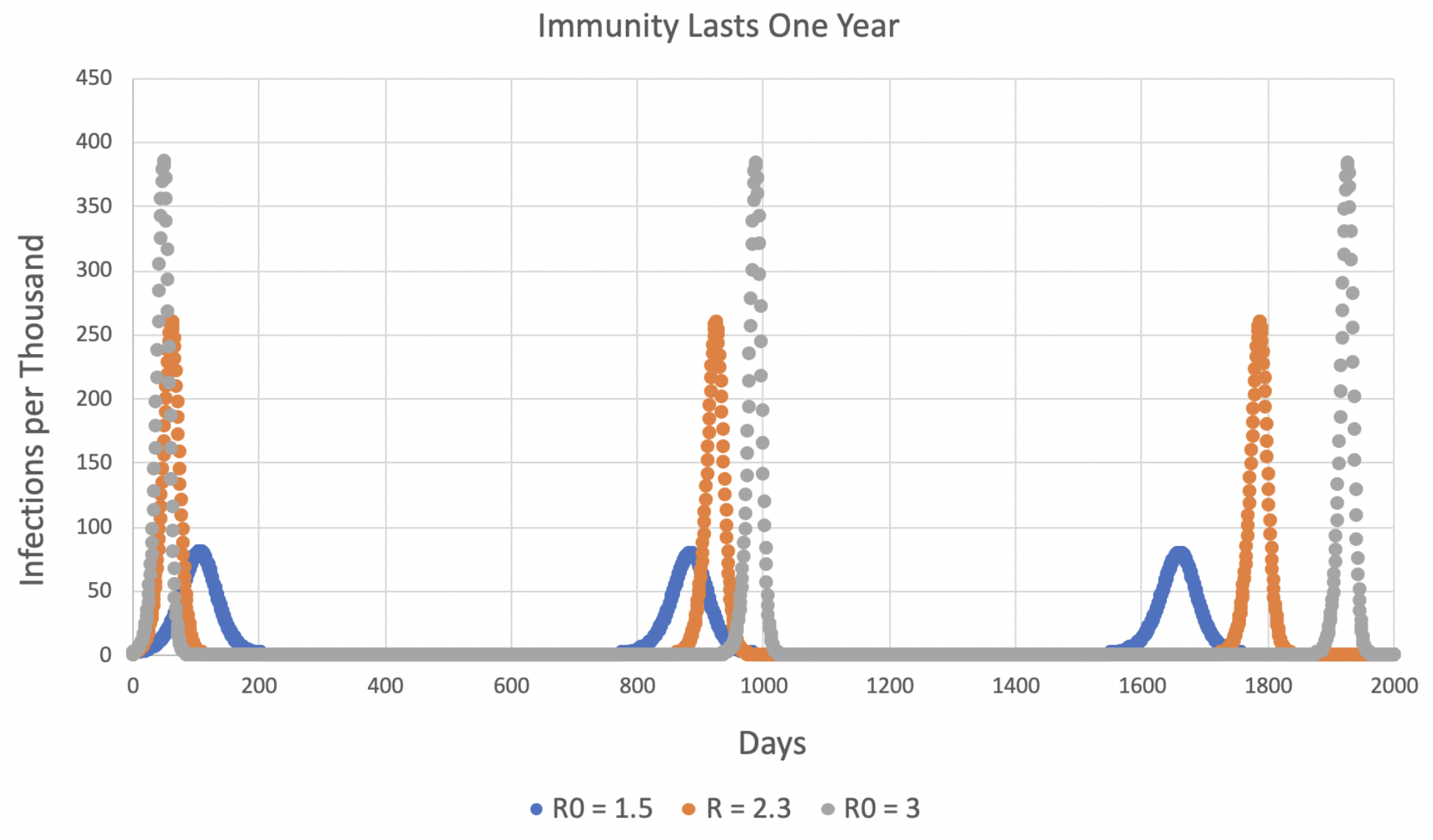
SEIRS model with 52 weeks (one year) of immunity. Note that the x-axis extends out to 2000 days (more than five years). See Fig 7.

**Fig. 7.**
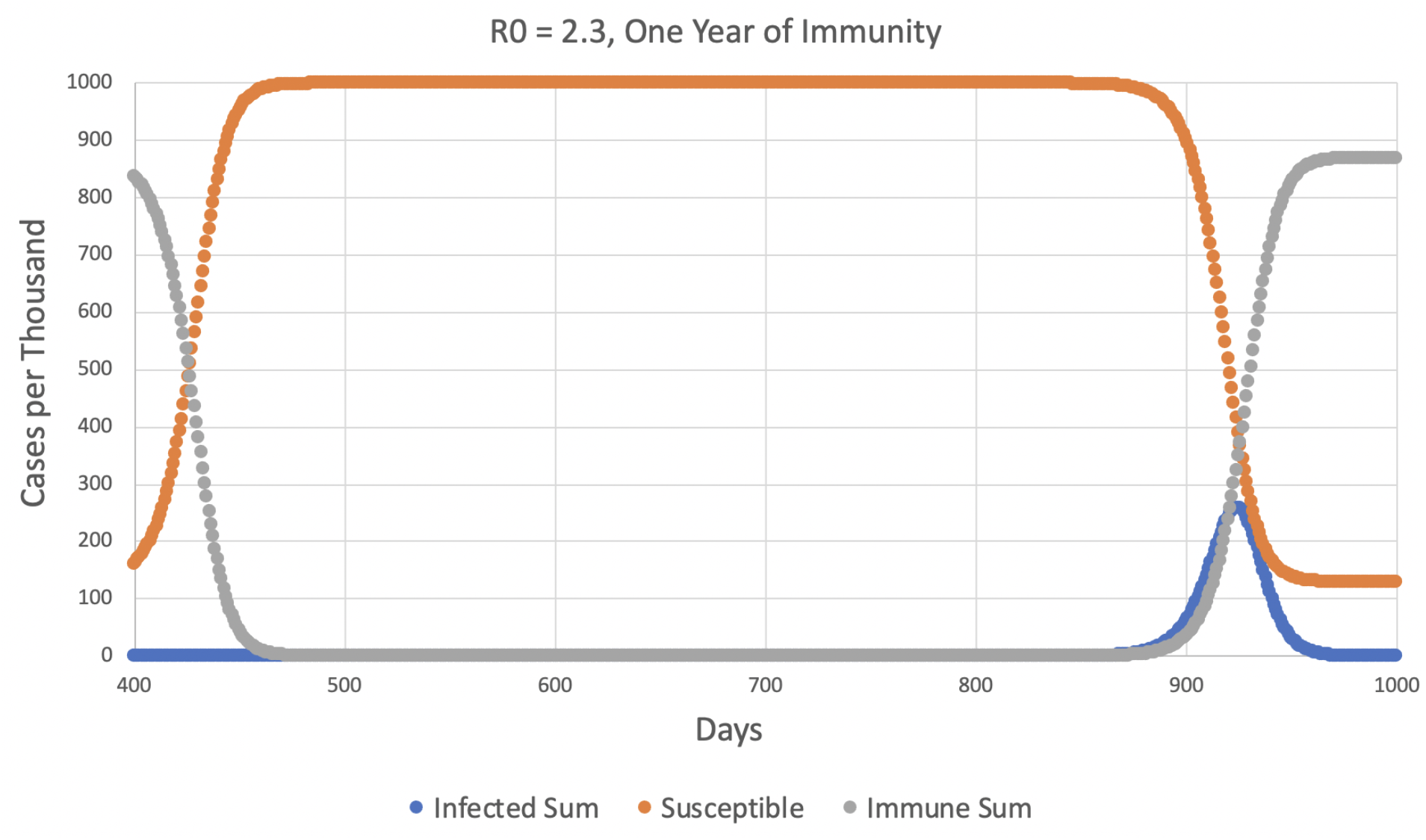
The portion of Fig. 6 from 400-1000 days using R_0_ = 2.3. Note how the decline in immune hosts due to the ending of their one year of immunity (gray) produces an increase in susceptible hosts (orange), and this in turn causes an increase in infected hosts (blue) after a lag of more than a year

**Fig. 8.**
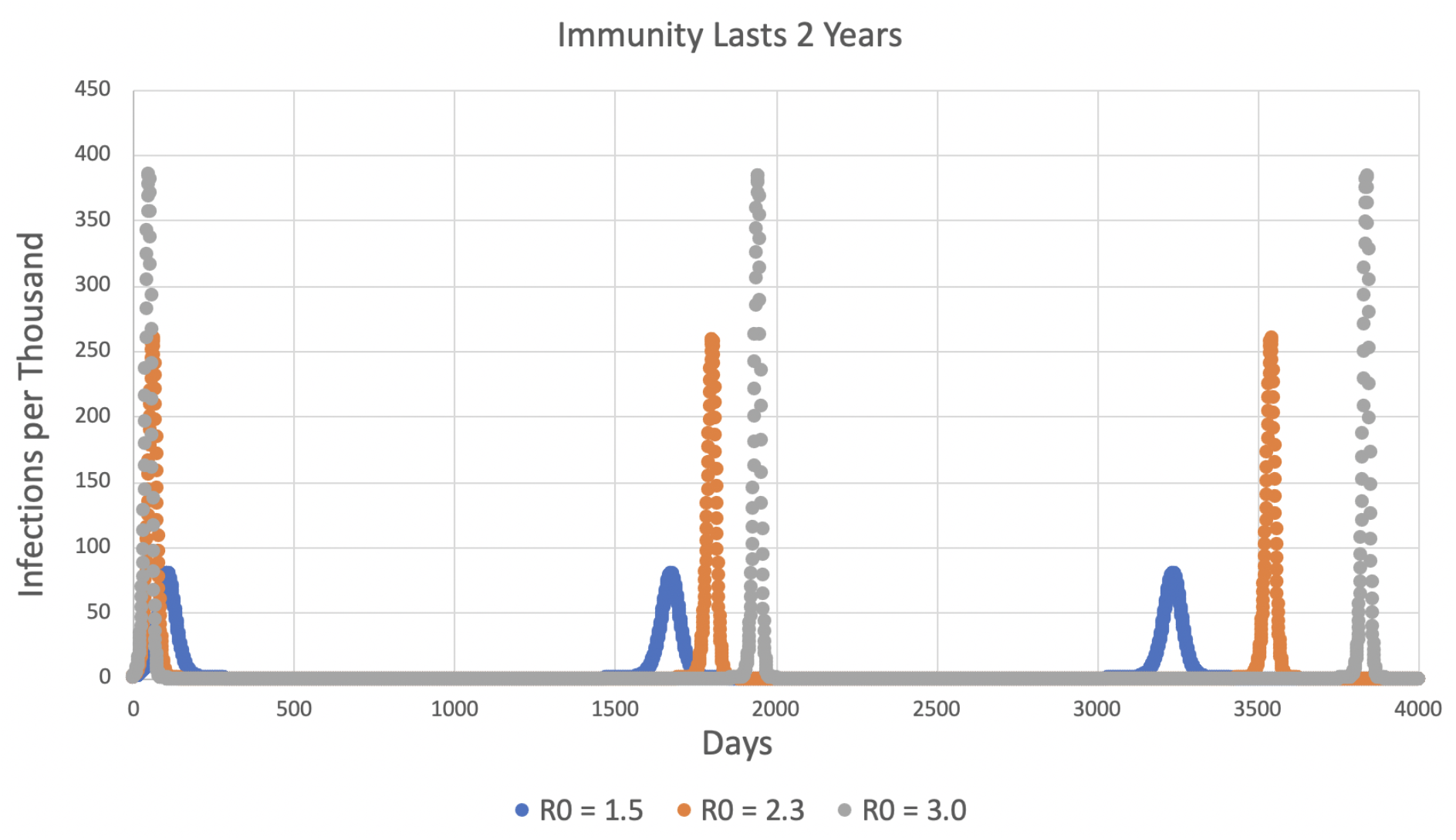
SEIRS model with 104 weeks (2 years) of immunity. X-axis extends to 4000 days (almost 11 years).

**Figure 9.**
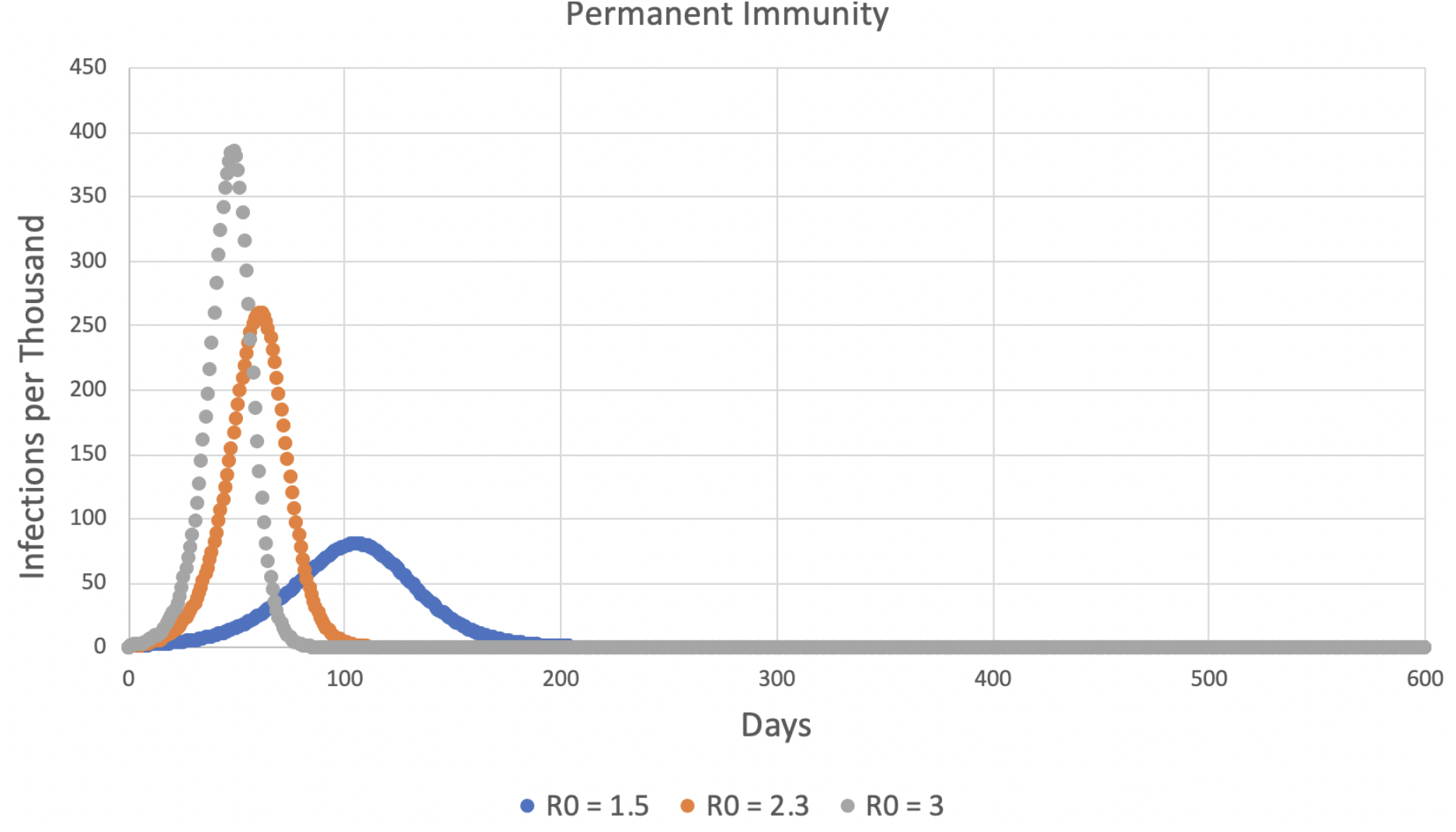
SEIR model with permanent immunity. The epidemic ends for all values of R_0_.

Figures 5, 6, and 8 show that long periods of immunity (but not permanent immunity) lead to outbreaks of disease at long intervals. This pattern is retained when immune fading is in effect, but the more complete the fading is, the more frequently the outbreaks become:

**Table 1.**
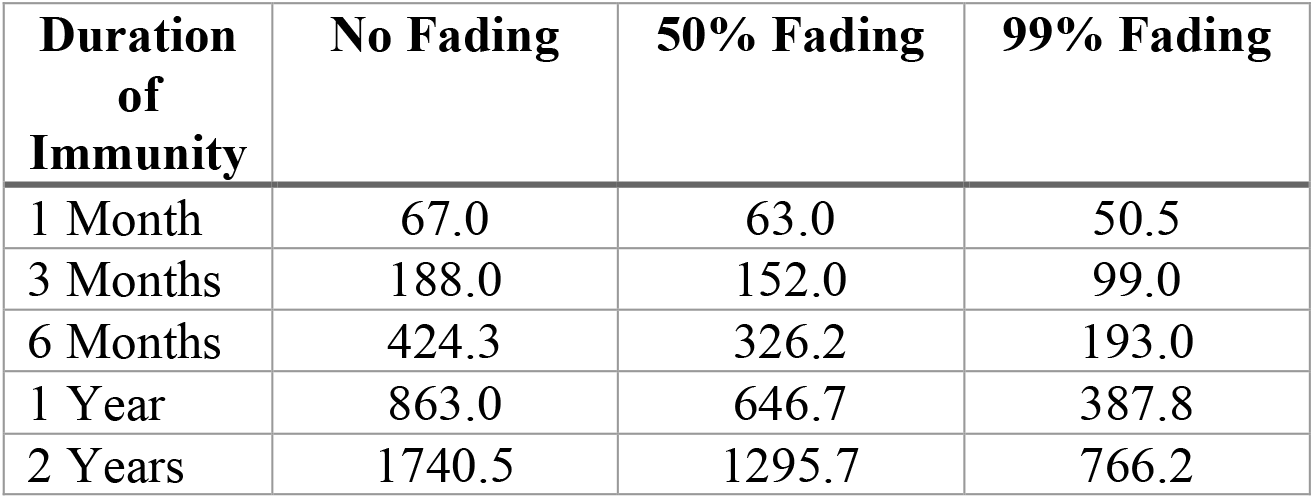
Time (in days) between outbreak peaks for 0%, 50%, and 99% immune fading for immune periods of various durations. R_0_ = 2.3 in all cases.

| Duration of Immunity | No Fading | 50% Fading | 99% Fading |
| --- | --- | --- | --- |
| 1 Month | 67.0 | 63.0 | 50.5 |
| 3 Months | 188.0 | 152.0 | 99.0 |
| 6 Months | 424.3 | 326.2 | 193.0 |
| 1 Year | 863.0 | 646.7 | 387.8 |
| 2 Years | 1740.5 | 1295.7 | 766.2 |

However, despite the more frequent outbreaks with greater immune fading, the peak number infected in each outbreak was virtually the same as when there was no fading.

For simulations *without* immune fading, the time between peaks is about 2.35-2.40 times the duration of immunity (e.g., 863 days for immunity that lasts 364 days).

## Discussion

This research produced both expected and unexpected results about the duration of immunity to a COVID-like disease.

On the expected results, Figs. 2 and 9 show that the SEIS and SEIR models as implemented on the spreadsheet produced the classic, anticipated results. Also, use of a higher R_0_ in the SEIRS models produced higher peaks, and more sweeping fluctuations that resisted settling down to an equilibrium value. For example, contrast the different R_0_ curves in Fig. 3.

The most *unexpected* outcome is that even relatively short periods of immunity could produce marked oscillations. Consider Fig. 3 again. While one month of immunity might be dismissed as epidemiologically unimportant, it was sufficient to create recurrent outbreaks of the epidemic approximately every 63 days for R_0_ = 3.0 and every 67 days for R_0_ = 2.3. The curve for R_0_ = 1.5 rapidly settled to an equilibrium. In Fig. 4, immunity lasting three months produced repeated outbreaks about every 200 days for R_0_ = 3.0, 187 days for R_0_ = 2.3, and about 180 days for R_0_ = 1.5.

Fig. 5 shows that for six months of immunity, all three values of R_0_ produced renewed outbreaks of the disease more than a year after the first outbreak had died away, and the new outbreaks were just as intense as the first one. The simulations using one and two years of immunity (Figs. 6, 7, and 8) continued this pattern of similar-sized outbreaks at long intervals (in Fig. 8, peaks about five years apart for R_0_ = 3.0). These strong, renewed outbreaks after immunity “times out” seem to be a robust feature of the model.

As mentioned previously, when there is no immune fading, the time separating the outbreaks appears to be about 2.4 times as long as the period of immunity.

Fig. 7 explains the timing of the outbreaks for one year of immunity. At the left side of the figure, immunity (the gray curve) is rapidly declining, and the susceptibles are going up as a result. By 500 days, 999.97 of 1000 organisms are susceptible and only 0.03 are immune. But infected hosts slowly increase over the next year, reaching 1.0 infected organism on day 860. From this point, the number of infections increases explosively.

“Immune fading,” in which immunity gradually fades away rather than persisting undiminished to the end of the immune period, does not change the height of the peaks, but it does make the outbreaks more frequent. The interpeak interval is mainly established by the ability of infected individuals to recover from very low numbers. Early release of immune individuals to the susceptible population keeps the infected population higher than it would be without fading. For example, in the simulations in which immunity lasted a year, the low point of the infected individuals between the first two peaks was 3.6 ⨯ 10^−20^ when there was no immune fading, but it was 5.3 ⨯ 10^−11^ with 50% fading and 6.7 ⨯ 10^−5^ with 99% fading. With higher numbers of infected individuals between peaks, recovery will be faster and the peaks will be closer together. Immunity where 99% of the individuals revert to susceptible before the immune period is finished can cut the interpeak time interval by more than half.

To summarize, while admittedly these simulations leave out much real-world complexity, they make it clear that the duration of immunity, *by itself*, can have a great influence on the course of the an epidemic. Shorter durations of immunity are not effective in ending the epidemic, but cause oscillations in disease prevalence. Longer periods of immunity can successfully suppress the disease for long periods, but are characterized by repeated outbreaks triggered by the expiration of immunity. Gradual “leakage” of individuals from the immune compartment to the susceptible compartment can markedly reduce the time period between outbreaks of the infection.

The duration of effective immunity is a key variable in the control of a COVID-19 epidemic.

## Data Availability

The MS is complete as it is. There are no extra data files

## Literature Cited

Britton, T., F. Ball, and P. Trapman. 2020. A mathematical model reveals the influence of population heterogeneity on herd immunity to SARS-CoV-2. Science 368:23 June 2020. No pages assigned yet. https://doi.org/10.1126/science.abc6810.

Kissler, S. M., C. Tedijanto, E. Goldstein, Y. H. Grad, and M. Lipsitch. 2020. Projecting the transmission dynamics of SARS-CoV-2 through the postpandemic period. Science 368: 860–868. https://doi.org/10.1126/science.abb5793.

Prompetchara, E., C. Ketloy, and T. Palaga. 2020. Immune responses in COVID-19 and potential vaccines: Lessons learned from SARS and MERS epidemic. Asian Pacific Journal of Allergy and Immunology 38: 1–9. https://doi.org/10.12932/AP-200220-0772.

